# Invisible pursuit: A scoping review of global policy for continuity of care of vulnerable infants under 6 months and their mothers in low- and middle-income countries

**DOI:** 10.1101/2024.12.06.24318470

**Authors:** Marie McGrath, Hedwig Deconinck, Stephanie V. Wrottesley, Marko Kerac, Tracey Smythe

**Author notes:** Corresponding author; Marie McGrath, (MM). **Contributorship** MM: Conceptualization, data curation, formal analysis, funding acquisition, project administration, supervision, investigation, methodology, visualisation; writing - original draft preparation, writing review and editing HD: Data curation, formal analysis, investigation, methodology, writing - original draft preparation; visualisation; writing - review and editing SW: Data curation, investigation, writing - review and editing MK: Conceptualization, methodology, writing - review and editing TS: Conceptualization, methodology, supervision, writing - review and editing.

## Abstract

Millions of infants worldwide are born vulnerable or become so in their first six months of life, which carries immediate and long-term risks. We conducted a scoping review to examine coherence of global policy content to inform national policy development.

We included policy guidance on vulnerable infants under six months and their mothers applicable to multiple settings in low- and middle-income countries. We applied JBI updated methodology and PRISMA-ScR guidance. We included English publications only with no time limit. We sourced documents through Google Scholar, personal contacts, databases, global networks and agency websites. The search was conducted from August 2023 to February 2024. From 91 identified documents, we prioritised 34 documents for review. We categorised policy into guidelines - WHO documents with evidence-based recommendations, and guidance - multi-source documents with implementation details. We investigated characteristics, vulnerability descriptors, and continuity of care. We consolidated 49 vulnerability descriptors into 28 vulnerability factors and four sub-groups. We did not assess policy quality or undertake formal policy analysis.

We found rich but inaccessible global policy guidance. Multiple terminologies create superficial differences and mask important ones. Growth appraisal was mostly limited to nutrition-oriented guidance and lacking in health-centric documents. Continuity of care was present in principle but lacked depth, especially for mothers. WHO policies are out of sync with each other and the latest evidence on mortality risk markers. Potential to leverage what exists needs WHO procedures to accommodate non-UN documents. There are immediate opportunities and necessity for cross-speciality policy cooperation. A living policy system to navigate evidence-to-policy process and manage policy interactions is warranted for this vulnerable group. Applying the WHO-INTEGRATE tool to policy contextualisation could help country-led adaptations, system-sensitive global support, and WHO methodological development. Ambition, investment and accountable action is urgently needed to secure coherent, evidenced policy for equitable and effective care.

## INTRODUCTION

Many infants are born vulnerable or become so in the first 6 months of life, leading to poor growth and development, increased risk of illness and death, and long-term limitations for their health, educational, and economic potential. (1, 2) The burden is high in low- and middle-income countries (LMICs), where an estimated 10.3 million (17.4%) infants under 6 months (u6m) are underweight, 9.2 million (15.5%) are wasted, and 11.8 million (19.9%) are stunted (3). Worldwide, 8.9 million infants (14.6%) are born with low birth weight (LBW) each year (4) who are at even greater and continued risk, especially those born too early or too small (premature/small-for-gestational age) (5).

Vulnerable infants u6m and their mothers present to national health and nutrition services in different ways, such as LBW infants in reproductive health services, malnourished infants in growth monitoring programmes and sick infants in outpatient health clinics. Their mothers may have their own physical, mental or social vulnerabilities that necessitate targeted support. Consequently, continuity of care, requiring access to the health services they need, delivered equitably and with quality, requires health systems to connect services within and across specialities (6).

WHO provides evidence-based guidelines (7) and other normative products that guide Member States on their public health decisions and actions (8). Examples applicable to vulnerable infants u6m and their mothers include WHO guidelines on malnutrition (9), maternal and newborn care (10), LBW and preterm infants (11), and Integrated Management of Childhood Illness (IMCI) (12, 13). WHO, and many other United Nations (UN), non-governmental (NGOs) and civil agencies, assist national authorities to implement these recommendations in their contexts. Initiatives include development of implementation guidance and direct involvement in national policy update processes as specialist advisors, for example. Such initiatives are underway for the updated 2023 WHO malnutrition guideline (14) and for follow-up care of small and sick newborns. Policies have varied update processes and timelines; an IMCI update is in progress. Increasingly, WHO is moving towards a living guidelines approach, enabling responsive updates to specific recommendations as evidence emerges (15).

Coherent national policy enables governments to deliver on strategic ambitions through collective action (16). This increases the potential for achieving inter-speciality continuity of care to the benefit of those accessing services and those working within them. National policymakers must consider new or updated WHO recommendations relative to what exists both at global and national level. Understanding the nature of global policy guidance can inform national policy reactions to new policy developments and those supporting their efforts on how best to do so (17). To help, we undertook this scoping review to investigate the nature and coherence of global policy that guide care of vulnerable infants u6m and their mothers. We chose the scoping review method as a recognised means of exploring and describing breadth and depth of complex, diverse literature to gain insights on implications for policy, research and practice (18-20). We examined global policy content to answer four questions:

1. What are the characteristics of global policy guidance related to vulnerable infants u6m and their mothers?
2. How is vulnerability in infants u6m and their mothers described?
3. How is continuity of care for vulnerable infants u6m and their mothers conceptualised?
4. What are emerging considerations for developing policy that addresses vulnerable infants u6m and their mothers?

## METHODS

### Approach

We used a published national policy scoping review protocol to set our objectives and align to national policy needs (16). We applied JBI updated methodology for scoping reviews (21) and used the Preferred Reporting Items for Systematic Reviews and Meta-Analysis extension for Scoping Reviews (PRISMA-ScR) to guide reporting (22) (S.Appendix 1). We used a ‘learning by doing’ approach to handle complexity, by using iterative, reflective learning to refine our thinking and adapt our processes (23). We shared the rationale, objectives, and invited collaboration with WHO staff working on related policy guidance initiatives and consultations. A practitioner lens informed decisions on how to categorise, prioritise, map and synthesise findings. We drew on our collective expertise and external reviewers to consider what is feasible and useful for practice-informed policy interpretation, implementation and development. A Reflexivity Statement is included in S.Appendix 2 and embedded throughout this paper’s narrative (24).

### Definitions, terminologies and frameworks

Definitions and terminologies used in this review, that build on WHO glossary of terms (25), is shown in Box 1. To enhance readability, we sub-categorise global policy guidance (hereafter referred to as policies) into three groups:

a. guidelines: WHO-specific documents with evidence-based recommendations
b. guidance: multi-source and multi-type documents detailing implementation, and
c. enabling documents: multi-source and multi-type documents that frame implementation

We use the term ‘vulnerable’ to encompass all definitions/descriptors/markers of risk for infants u6m and mothers. For the sake of clarity and brevity, the term “mother” is used to denote “mother or principal caregiver,” with the understanding that these roles may not always be synonymous in practice. We apply the term/derivatives “malnutrition” to encompass wasting and oedematous malnutrition. ‘Low anthropometry’ refers to single measurements. “Poor growth” refers to sequential weight/mid-upper arm circumference (MUAC)/anthropometric index measures. We applied the WHO’s continuity of care definition (25).

We used the MAMI Care Pathway Package as a starting framework to appraise to what extent global policy guidance supports continuity of care for vulnerable mother-infant pairs (26) (S.Appendix 3). This approach was collectively developed by the MAMI Global Network (27) to support continuity of care for small and nutritionally at-risk infants u6m and their mothers (MAMI) within health systems.

#### Box 1

**Definitions, classifications and terminology used in this review**

**Policies** include normative WHO guidelines and related products and non-WHO implementation or programme guidance, briefs, training guides, manuals, action plans, strategies, and frameworks related to the care of vulnerable infants u6m and their mothers. Sources include United Nations (UN) agencies, professional networks, non-governmental organisations (NGOs), and civil society.

**A guideline** is any information product developed by WHO that contains evidenced recommendations for clinical practice or public health policy. Recommendations are statements designed to help end-users make informed decisions on whether, when and how to undertake specific actions such as clinical interventions, diagnostic tests or public health measures, with the aim of achieving the best possible individual or collective health outcomes.

**A guidance** is any information product that that describes how to support implementation. This includes implementation or operational guidance and manuals. It may be produced by WHO or other entities.

**Enabling documents** include frameworks, action plans, strategy papers, and training materials that support quality implementation, developing competencies, achieving set goals, improving quality, strengthening political commitment, creating opportunities to accelerate spread, and sustainable scale-up.

**Vulnerable mother-infant pairs** includes but is not limited to infants from birth to 6 months of age (u6m), including newborns born with low birth weight (LBW) or born premature or small-for gestational-age; infants with wasting or nutritional oedema (acute malnutrition), stunting, or underweight; infants with acute or chronic illness and disability or other growth and development concerns; and infants of mothers who are vulnerable because of poor physical or mental health or nutritional or social conditions.

**Care**, for the purpose of this review, covers physical and mental health and nutrition care for infants u6m and physical and mental health, nutrition, and social care for their mothers, across inpatient, outpatient and community settings.

**Continuity of care** is used to indicate one or more of the following attributes of care: 1) provision of services coordinated across levels of care–primary care and referral facilities, across settings and providers; 2) provision of care throughout the life-cycle; 3) care that continues uninterrupted until the resolution of an episode of disease or risk; and 4) coherent and interconnected healthcare events over time and consistent with people’s health needs and preferences. We considered quality, respectful care an inherent component of all attributes for person-centred care.

**Person-centred care** consciously adopts the perspectives of individuals, families, and communities as participants in and beneficiaries of trusted health systems. Such perspectives include respect for persons’ values, preferences, and expressed needs regarding coordination and integration of care; information, communication, and education; physical comfort, emotional support, and alleviation of fear and anxiety; involvement of family and friends; and transition and continuity.

### Eligibility criteria

We used the ‘population, concept, and context’ (PCC) mnemonic to create eligibility criteria (21).

#### Population

Newborns and infants u6m who were born or became vulnerable and their mothers, in LMICs.

#### Concept

Documents that provided global policy guidance on infant health and nutrition, child development, maternal physical and mental health, nutrition, and childcare and feeding practices. For example, support for breastfeeding, clinical and public health interventions for vulnerable infants u6m and their mothers from birth, active growth monitoring, food or supplementation interventions, and continuity of care across services and over time.

#### Context

We considered all settings and contexts for the care of vulnerable infants u6m and their mothers in LMICs, including but not limited to primary, secondary and tertiary healthcare that may involve inpatient, outpatient and community-based services, including within the home.

We excluded regional or national policies as we sought policy guidance applicable to multiple settings and contexts (for exceptions that emerged, see Boundaries and Limitations). Due to capacity constraints, we included documents in English only. We did not set a time limit for policy inclusion.

### Information sources

We sourced publicly available documents through Google Scholar, personal researcher files (MM, HD) professional contacts, global health and nutrition networks and UN/NGO websites from August 2023 to end of February 2024. The Google Scholar Search was conducted in January 2024. We sourced data from two publicly available databases – the WHO Institutional Repository for Information Sharing (IRIS) (28) and UNICEF data repository (29), six international agency websites and via direct contacts. We hand-searched document references and snowballed to further source content. We followed up with 42 individuals identified through personal contacts and open calls to communities of practice (MAMI Global Network, CORE Group, Global Nutrition Cluster). The professional and agency affiliations of contacted individuals is included in S.Appendix 4. The policy documents database (in Excel) is available upon request. Documents we included in this review are available at the online Tableau Public platform (30).

### Search strategy

We explored global policy from multiple sources relating to the care of vulnerable infants u6m and their mothers. We first searched for global policy guidance related to health, nutrition, child development and care for vulnerable infants u6m in English, without date restrictions. We considered source, official categorisation and nature of their content. Three researchers were involved in the process (MM, HD, SW). The search strategy is included in S.Appendix 5.

### Selection of sources

We examined identified policy documents in five steps (Figure 1) as follows:

**Figure 1:**
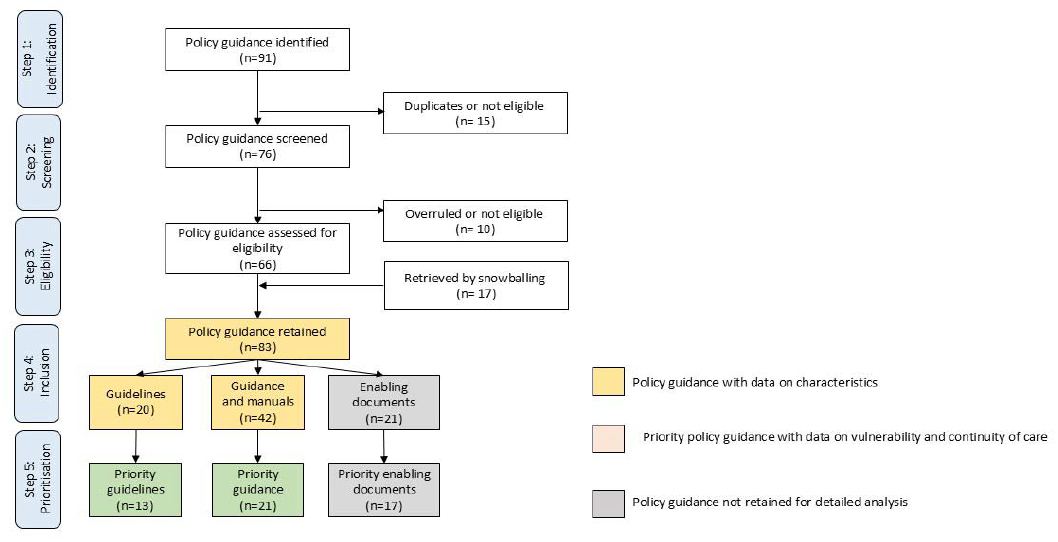
Flow chart on selection of global policy for vulnerable infants u6m and their mothers.

#### Identification

Compiled citations from grey literature searches and key contacts in an Excel spreadsheet and removal of duplicates (*MM framing, SW compile, MM and HD review & confirm*)

#### Screening

Screened titles and summaries to exclude irrelevant policy, resolution of inconsistencies and decision-making through team discussions (*MM, HD, SW, confer & confirm*)

#### Eligibility

Screened full texts, removal of discrepancies or outdated policy and addition of missed documents *(SW screen, MM and HD review & confirm)*

#### Inclusion

Classified retained global policy into a) guidelines b) guidance and c) enabling documents *(SV & HD compile, MM review & confirm)*

#### Prioritisation

Identified policy that specifically addressed vulnerability in infants u6m, from that addressing healthy newborns or general, promotive infant or maternal health *(MM & HD confer & confirm)*

Data screening and selection to source priority policies involved three phases within Steps 4 to 5 (Figure 1):

- In Phase 1, we appraised characteristics of 83 retained policy documents: 20 guidelines, 42 guidance and 21 enabling documents. We excluded 21 enabling documents.
- In Phase 2, we conducted more in-depth exploration of the nature and coverage of vulnerability factors in 62 retained guidelines and guidance. We identified 34 priority policies.
- In Phase 3, we appraised 34 priority policy for coverage and content regarding vulnerability factors and continuity of care.

In May 2025, we further appraised data in reaction to newly published evidence mortality risk markers for infants and children (31).

### Data Charting Process

Data was charted using customised forms (MS Excel). Through the process, we iteratively organized information and adjusted our guiding frameworks accordingly.

#### Characteristics

In Phase 1, HD extracted key characteristics for each study, using a custom form (MS Excel). Data included publication year, URL, source, title, type, aim, topic, target audience, target population, infant descriptors, maternal descriptors, sector, level of care, care service, age timeline, infant risks, maternal risks, interventions, and reviewer notes. MM independently reviewed data extracted. Discrepancies were discussed and resolved by MM, HD and SW.

We extracted content on speciality or discipline, e.g., child health, child nutrition, child development, maternal physical health, maternal mental health, maternal nutrition, reproductive health, neonatal health) and on sector/level (e.g., tertiary, secondary, primary, or community care).

We extracted content on interventions e.g., active case finding or screening, health assessment, breastfeeding or non-breastfeeding assessment, breastfeeding or non-breastfeeding support, clinical care, nurturing care, Kangaroo mother care, early childhood development, crying and sleep counselling, mental health counselling, social support, follow-up visits, home visits, family involvement, and continuity of care.

We extracted content on condition, such as health problem, disease, disorder, injury, disability, or circumstance.

#### Vulnerability

In Phase 2, for infant vulnerability, we explored for content on preterm, LBW, small-for-gestational age, low anthropometry, poor growth, feeding problem, metabolic problem, excessive crying, disability, and acute or chronic illness. For maternal vulnerability, we looked for nutritional risk, impaired breastfeeding conditions, physical health, maternal mental health, and mothers who are multipara, primipara, adolescent, absent, or dead.

We identified 49 vulnerability factors in policy documents. Since some were very similar, we merged these into 28 vulnerability factors. To help navigate data and findings, we categorised the 28 vulnerability factors into four sub-groups: 1) poor birth outcomes (includes small and sick newborns),2) low anthropometry or poor infant growth 3) other infant risk factors and 4) other maternal risk factors.

In Phase 3, we categorised and mapped 24 vulnerability factors across 4 sub-groups by policy type. We divided vulnerability factors at the median (equal or above [≥] the 50% threshold) to determine which factors were most frequently covered. We examined coverage of three mortality risk markers – low weight-for-age z score (WAZ), history of LBW or preterm birth, and not breastfeeding (31).

#### Continuity of care

From our baseline MAMI Care Pathway framework, we iteratively identified 11 care dimensions, specifically care across time, services and levels of care; integrated care pathway; comprehensive person-centred care; early childhood development; mother, father, family support; community participation; embeddedness; local health system support; monitoring and evaluation of services; wider multisectoral support; and organisational capacities, including resilience. A dimension fully or partially addressed was classified as ‘applied’. We mapped the relevant condition for each. Finally, we mapped which care dimensions and conditions were covered by each policy.

### Data items

Data items used to appraise sources in Phases 1-3 and to chart data is available in S.Appendix 6. The variables comprised characteristics, vulnerability descriptors and continuity of care dimensions. We explored applying a life-cycle dimension but found it oversimplified and risked misrepresentation of content (e.g., a guidance may state a greater age-range than was detailed in guidance content). We did not appraise the quality of guidance but applied components of the AGREE II (Appraisal of Guidelines for Research and Evaluation) checklist relevant to policy development processes (scope and purpose, stakeholder involvement, and whether a procedure for update provided) (32, 33).

### Synthesis of Results

Results were narratively synthesised supported by summary tables and supplementary appendixes.

### Sharing of Results

Through 2024 to current date, data and scoping findings were shared with WHO and UNICEF, via the MAMI Global Network (co-chaired by MM and MK), in a report (34), and with policymakers in Ethiopia undertaking national malnutrition policy updates (MM).

## RESULTS

### Search results

Initial searches sourced 67 potential documents, 78% (n=52) from databases, 18% (n=12) from agency websites and 4% (n=3) from Google Scholar. Most database documents were sourced from the WHO Iris Repository (85%, n=44). Over half of website sourced documents (n=7, 55%) were located from six NGO websites, one-third direct from the WHO website (36%, n=4) and 9% (n=1) from the UNICEF website. Of website sourced documents (n=12), three agencies were hosts to six collectively produced policy documents - Quality of Care Network, WHO host; Healthy Mothers Healthy Babies Consortium, UNICEF host; and MAMI Global Network, ENN host. A further 24 documents were sourced for appraisal through snowballing references, contacts and researcher personal files to yield a total of 91 eligible policy documents. From these we shortlisted and characterised sixty-two policies, which consisted of 20 guidelines (11, 35-53), and 42 guidance (13, 26, 54-93). From these 34 priority policies were eligible for review, which consisted of 13 guidelines (11, 35-46) and 21 guidance documents (13, 26, 54-72).

### Characteristics

#### Source

All 13 guideline documents were published by WHO Geneva, except one from the regional Pan American Health Organization (39). Twenty-one guidance documents were produced by a wide range of organisations, led/co-led by UN agencies (WHO, n=12, 57%; UNICEF (n=5, 24%), NGOs (n=2, 10%), specialist agencies/foundations (n=2, 10%), academics (n=1, 5%), and one network involving multiple agencies/individuals (5%).

One guideline document had a direct WHO operational guidance counterpart (36, 55). While other guideline documents did not have a direct counterpart, we identified content consistent with guideline recommendations both within and beyond UN-authored guidance, e.g., for early childhood development (40, 62), LBW/preterm (11, 57, 65) and malnutrition/poor growth (26, 35). In some cases, such guidance preceded the WHO recommendations that it provides relatable implementation details on (26, 35).

#### Publication dates and updates

Publication dates ranged from 2012 to 2023 for guideline documents and from 2003 to 2022 for guidance documents. Two guideline documents (15%) specified an update date, six make no reference to update (19%). Over one-third (n=5, 38%) recorded a less specific intent, such as update with emerging evidence, in line with other guidance developments, or state they may consider it in the future. Six guideline documents (46%) included a named or departmental contact. Amongst guidance documents, only two documents (10%) specified an update date (year) and one indicated intent. Eleven (52%) provided a contact person or department.

#### Condition

Guideline documents covered four conditions related to newborn care, six on illness, one on low anthropometry and poor growth, one on breastfeeding and one on early childhood development. Guidance documents covered five conditions on newborn care, seven on illness, two on preterm and LBW, two on poor growth and malnutrition, one on breastfeeding, one on feeding difficulties, one on perinatal mental health, and one on reproductive health.

Some guideline documents targeted a single condition (e.g., tuberculosis) (36-38, 42, 43, 45) or a set of conditions (e.g., common childhood illness, poor growth and development) (35, 46). Others targeted specific vulnerability profiles, e.g., LBW newborns (11, 39, 44) or embed content specific to them within a broader condition/population policy (40, 41). Care delivery point ranged from inpatient facilities (e.g., resuscitation) to community (e.g., outpatient facilities) and home level (e.g., breastfeeding support).

Guidance documents targeted single conditions (e.g., cerebral palsy) (55, 70) or sets of conditions (e.g., birth impairments) (13, 66, 68), interventions (e.g., Kangaroo mother care, early childhood development) (61-64) and profiles of vulnerability (e.g., small sick infants, preterm infants) (65, 67, 71). Some targeted multiple at-risk infants u6m (26, 62) or sub-sets of these, e.g., preterm/LBW newborns (57). Some guidance documents focused on quality of care and behaviours or factors that influence vulnerability, e.g., feeding difficulties, maternal mental health (56, 59-61). One guidance related to service integration (58).

#### Target audience

Across guideline documents, the target audience was broad and diverse across specialities or sub-specialities within health and nutrition; system levels and professional remit, most often policymakers, programmers, health workers, trainers; and institutions, typically government, UN, NGOs, academics and funders. Guidance documents were more oriented toward health practitioners, clinicians or service planners; one-third (n=5, 36%) specified policymakers as a target.

#### Target population

All 13 guideline documents targeted infants u6m, eight (61%) incorporated aspects related to mothers (11, 35, 37-41, 44). Twelve guideline documents (57%) were applicable from birth, of which three were specific to the early postnatal period (43-45). One guideline document from birth limited one recommendation (nutritional supplementation) to infants from 1 month of age (35).

All 21 guidance documents covered infants u6m or a sub-set, half (n=11, 52%) included mothers of which one targeted perinatal mental health (56). Nine (21%) guidance documents related to other aspects of service provision, e.g., integration of services, monitoring and evaluation, resources and equipment, and quality of care.

#### Vulnerability descriptor for infants

Across all policies, many descriptors were identified for infant vulnerability according to topic, condition and speciality, including healthy newborn, healthy infant, vulnerable newborn, born preterm, LBW, small, small sick, (non)breastfed, developed wasting or acute malnutrition, underweight, stunting, growth faltering, at risk of poor growth and development, feeding problem, excessive crying, physical disability or developmental delay, and with acute or chronic illness.

#### Vulnerability descriptor for mothers

As with infants, many descriptors were identified for maternal vulnerability including healthy, sick, mental health issue, malnourished, adolescent, prenatal, perinatal, postnatal issues, (not)lactating, absent, multipara, primipara, absent, dead. Female-headed households was not identified as a vulnerability in any policies.

#### Sector

Guideline development were led by specialists in child health (n=9, 69%), involving sub-specialities (neonatal care (n=6, 46%), urgent care (n=3, 23%), disease specialists (n=3, 23%), maternal health (n=2, 15%)); specialists in nutrition (breastfeeding, malnutrition) (n=2, 15%), and in early childhood development (n=1, 8%). Guidance development were led by specialists in newborn care (n=6, 29%), nutrition/feeding (n=4), 19%), urgent care (n=1, 5%), illness management (n=6, 29%), disability (n=2, 10%), maternal mental health (n=1, 5%), reproductive health (n=1, 5%) and inter-sector/disciplines (n=4, 19%).

Some guideline documents cross-referenced to others, for example, 2023 malnutrition guidelines defer to neonatal guidance for nutritional supplementation in infants under 1 month of age (47). Others embedded details of recommendations within their document, e.g., the tuberculosis guideline document specifies anthropometric criteria for severe acute malnutrition (36); the 2023 malnutrition guideline (35) embeds detailed elements of IMCI (13). WHO guideline documents did not cross-reference to non-UN produced documents.

#### Level of care

About one-third of guideline documents (n=5, 38%) encompassed all levels of care, one-third specified facility-based care (n-5, 38%) and one-quarter focused on primary/community care (n=3, 23%). More than half (n=11, 52%) of guidance documents covered care at primary and secondary levels, 19% (n=4) mainly covered inpatient or specialist care, and three were more generalisable across all care levels (14%).

#### Care service

Guideline documents covered neonatal health, child health, maternal health, infant and young child feeding (breastfeeding) and malnutrition treatment services, to widely varying extents. This was influenced by the number and nature of recommendations and the extent of elaboration in good practice statements and notes.

Guidance documents had similar service scope and variation to guideline documents. However they included more specific and detailed sub-speciality or sub-group content, e.g., for early childhood development, children with disability, feeding difficulties (57, 60, 62, 70) and more cross-sectoral comprehensive, coverage of care (26, 65, 66).

### Vulnerability factors

Coverage of 28 consolidated vulnerability factors across the 34 policies reviewed is shown in Table 1 and mapped by policy document in S.Appendix 7. Sixteen factors were specific to the infant and 12 factors were specific to the mother.

**Table 1.** Vulnerability factors mapped across 34 policy documents by policy guidance type.

|  | Guideline documents* |  | Guidance documents |  | All policy documents |  |
| --- | --- | --- | --- | --- | --- | --- |
| **Vulnerability factors (n=28)** | N=13 | (%) | N=21 | (%) | N=34 | (%) |
| **Poor birth outcomes** |  |  |  |  |  |  |
| Congenital illness | 7 | (54) | 15 | (71) | 22 | (65) |
| Low birth weight (LBW)** | 7 | (54) | 14 | (67) | 21 | (62) |
| Preterm** | 7 | (54) | 13 | (62) | 20 | (59) |
| Disability or congenital abnormality | 5 | (38) | 12 | (57) | 17 | (50) |
| Birth trauma or complications | 5 | (38) | 7 | (33) | 12 | (35) |
| Preterm morbidity | 4 | (31) | 11 | (52) | 15 | (44) |
| Small-for-gestational age | 3 | (23) | 5 | (24) | 8 | (24) |
| **Low anthropometry or poor growth** |  |  |  |  |  |  |
| Low anthropometry (low WAZ**, low WLZ or low MUAC) | 3 | (23) | 7 | (33) | 9 | (26) |
| Poor growth (weight loss, stagnant or insufficient weight gain) | 3 | (23) | 12 | (57) | 15 | (44) |
| Nutritional oedema | 3 | (23) | 6 | (29) | 9 | (26) |
| **Risk factors related to the infant** |  |  |  |  |  |  |
| Breastfeeding difficulties | 7 | (54) | 14 | (67) | 21 | (62) |
| Illness | 7 | (54) | 11 | (52) | 18 | (53) |
| Not breastfed** | 4 | (31) | 13 | (62) | 17 | (50) |
| Neurodevelopment concerns | 4 | (31) | 11 | (52) | 15 | (44) |
| Hospitalisation history | 1 | (8) | 5 | (24) | 6 | (18) |
| Mental health | 0 | (0) | 3 | (14) | 3 | (9) |
| Risk factors related to the mother |  |  |  |  |  |  |
| Physical health | 6 | (46) | 15 | (71) | 21 | (62) |
| Breastfeeding conditions | 5 | (38) | 12 | (57) | 17 | (50) |
| Mental health | 5 | (38) | 11 | (52) | 16 | (47) |
| Social or contextual factors affecting care and feeding practices | 3 | (23) | 11 | (52) | 14 | (41) |
| Birth complication | 3 | (23) | 6 | (29) | 9 | (26) |
| Adolescent | 2 | (15) | 7 | (33) | 9 | (26) |
| Absent or died | 2 | (15) | 5 | (24) | 7 | (21) |
| Multipara | 1 | (8) | 6 | (29) | 7 | (21) |
| Primipara | 1 | (8) | 2 | (10) | 3 | (9) |
| Anaemia | 0 | (0) | 3 | (14) | 3 | (9) |
| Birth spacing | 0 | (0) | 2 | (10) | 2 | (6) |
| Low MUAC | 0 | (0) | 2 | (10) | 2 | (6) |

\*In descending order by frequency in guideline documents
\*\*Risk markers for increased infant u6m mortality identified in published analysis (31).

In guideline documents, factors mentioned most frequently (≥50%) were associated with poor birth outcomes (congenital illness, LBW, preterm), breastfeeding difficulties, and illness of infants (Table 2). Factors less frequently mentioned (40-50%) related to mothers’ physical health. In guidance, factors most frequently mentioned (≥50%) were associated with small and sick newborns (poor growth, breastfeeding difficulties, no breastfeeding, and infant illness) and mother wellbeing (physical health, breastfeeding conditions, mental health, and social or contextual factors).

**Table 2.** Vulnerability factors present in at least half of 34 policy documents.

| Category of vulnerability | Guideline documents<br>($>50\%$ coverage) | Guidance documents<br>($>50\%$ coverage) |
| --- | --- | --- |
| Poor birth outcomes | Congenital illness<br>Low birth weight<br>Preterm | Congenital illness<br>Low birth weight (LBW)<br>Preterm<br>Disability and/or congenital abnormality<br>Preterm morbidity |
| Low anthropometry or poor growth | No | Poor ponderal growth |
| Risk factors related to the infant | Breastfeeding difficulties<br>Illness | Breastfeeding difficulties<br>Not breastfed<br>Illness<br>Neurodevelopment concerns |
| Risk factors related to the mother | No | Physical health<br>Breastfeeding conditions<br>Mental health<br>Social or contextual factors |

#### Anthropometry and growth

Around one-third of guideline documents included low anthropometry or poor growth as a vulnerability factor (n=4, 31%) (35, 36, 39, 46). Of these, three included low anthropometry - one specified low WAZ (35), two specified low weight-for-length z score (WLZ) (36, 46).

Coverage was higher in guidance documents; over half included low anthropometry or poor growth (n=13, 62%). Of these, seven (33%) included low anthropometry - two included low WAZ for 0-6 months (26, 62), two IMCI guidance applied WAZ to 0-2 months and WLZ to 2-6m (13, 66), three WHO guidance applied WLZ criteria (54, 55, 69), one applied MUAC from 6 weeks to 6 months (35) and one applied MUAC from birth (26). Anthropometric or growth vulnerability were not included in guidance documents on disability, (68, 70), follow-up of at-risk newborns including community care, (65), feeding support (57) and peri-natal mental health (56).

#### Feeding

Breastfeeding difficulties (related to the infant) or breastfeeding conditions (related to the mother) were commonly included in policies to identify infant vulnerability. Not being breastfed was included more often in guidance documents (n=13, 65%) than in guidelines (n=4, 31%).

#### Maternal

Maternal vulnerability (e.g., breastfeeding conditions, mental and physical health, and social or contextual factors affecting care and feeding practices) were referred to less often in guideline documents (n=10, 77%) compared to guidance documents (n=17, 85%). Coverage in guideline documents was highest in those related to early postnatal care. None of the guideline documents and two guidance documents (26, 62) included mother’s MUAC, while other anthropometric indicators (e.g., weight) were absent from all. Adolescent motherhood was referred to in two guideline documents (15%) and one-third of guidance documents (n=7, 33%). Other less commonly identified vulnerabilities included birth complications and short birth spacing.

#### Illness

Guideline documents covered four conditions related to newborn care, six on illness, one on low anthropometry and poor growth, one on breastfeeding and one on early childhood development. Guidance documents covered five conditions on newborn care, seven on illness, two on preterm and LBW, two on poor growth and malnutrition, one on breastfeeding, one on feeding difficulties, one on perinatal mental health, and one on reproductive health.

With the exception of one guideline document on common childhood conditions (46), LBW/prematurity was not included as a vulnerability factor in clinical (sick infant care) guideline documents. All clinical guidance documents included LBW/prematurity as a vulnerability, except for tuberculosis operational guidance (55). Overall, there was little policy content specific to infants who were born small-for-gestational age.

#### Combination of risk markers

Individual and combined coverage of three risk markers for infant mortality (low WAZ, not-breastfed, LBW/premature) was low across policies (31). Only one guideline document (35) and two guidance documents (62) include all three risk markers for infants up to six months of age. LBW/prematurity was not specified as a risk factor in IMCI guidance and low WAZ is recommended for infants 0 to 2 months only (13, 66). Clinically-oriented guidance did not include WAZ; one WHO guidance included WAZ reference tables but did not specify weight assessment and appraisal as part of case management (69).

### Continuity of care

Table 3 maps each policy document across 11 dimensions of continuity of care and by condition. Coverage of dimensions varied greatly across guidelines and guidance. Table 4 summarizes the coverage of continuity of care by policy type and provides a summary appraisal of findings for each dimension. There was high coverage across several policy documents, some that were condition-specific, such as on perinatal mental health (56), some population-specific, such as hospital care for small, sick and preterm newborns (61). However, we found that high coverage often reflected intent but did not equate with in-depth content to guide on how to achieve this. Implementation details more often related to management continuity for conditions within services, with limited coverage of operational and wider multi-sectoral aspects of continuity of care.

**Table 3.**
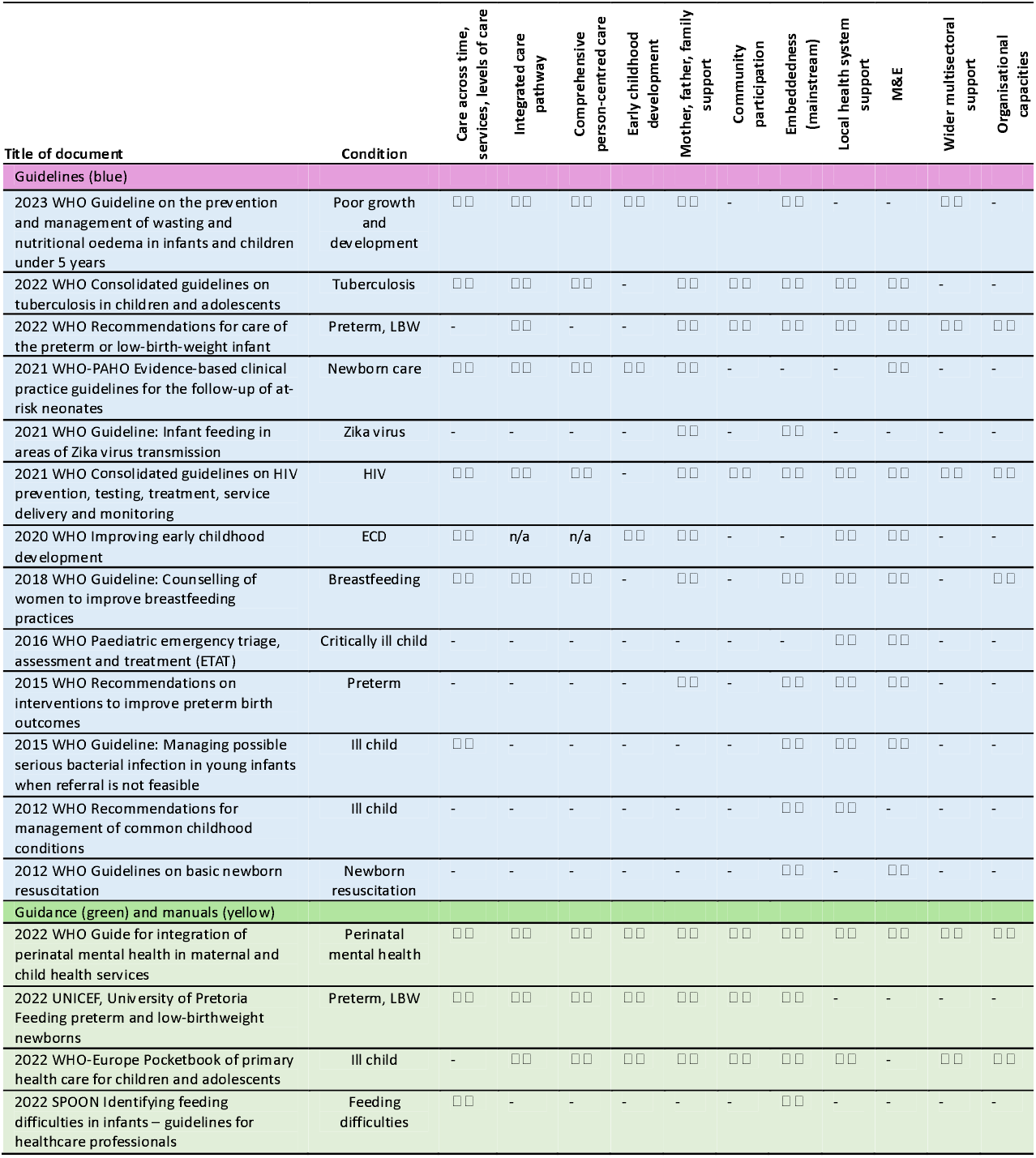
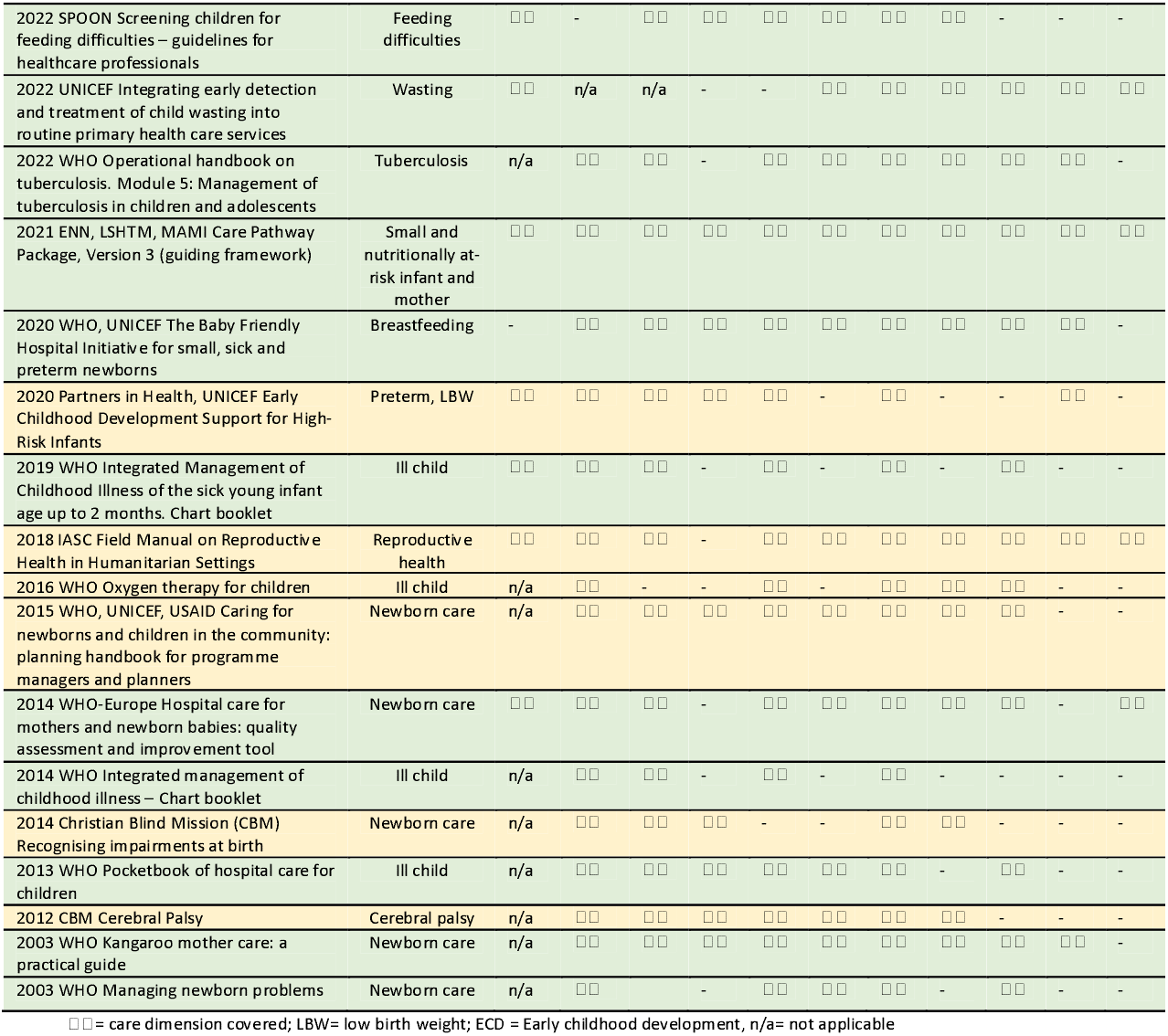
Coverage of continuity of care dimensions and related conditions across 34 policy documents.

**Table 4.** Coverage and appraisal of continuity of care across 34 policy documents by policy type.

| Dimensions of care | Guideline documents<br>(n=13) | Guidance documents<br>(n=21) | Summary appraisal* |
| --- | --- | --- | --- |
| Care across time, services and levels of care | 7 | 10 | Gaps in continuity across time, primarily for follow up after discharge from a particular service or when infants 'age out' of the service without adequate referral mechanisms. Also gaps in continuity between services and levels of care, e.g. linking facility- and community-based care. |
| Integrated care pathway | 6 | 18 | Strong focus on addressing one or more condition(s), without integration into a broader care pathway that assesses and responds to maternal and infant risks and monitors progress/ links to follow-up care, particularly at other levels of the health system (primary care/outpatient/community levels). |
| Comprehensive person-centred care | 5 | 17 | Guidelines centred on the condition rather than the person (mother, infant), and there was a lack of comprehensive assessment and care for both mother and infant together. |
| Early childhood development | 3 | 12 | Limited contextualisation within a framework that supports early child development, with a focus on identification and management of one or more condition(s) rather than on a holistic approach to ensuring optimal growth and development. |
| Mother, father, family support | 9 | 18 | Primary focus on supporting the mother in feeding and care practices for the infant, including targeted counselling for mothers and other caregivers/ family members in some cases. Lack inclusion of broader aspects of support, particularly for the mother's own health and social circumstances. |
| Community participation | 3 | 15 | Limited aspects of community participation in care/organisation of care, particularly for guidelines. |
| Integration, embeddedness (mainstream) | 10 | 21 | Focus on mainstreaming of guidance into routine care but lacking detail on how to embed services across levels of the health system (particularly primary care/outpatient/community). |
| Local health system support | 9 | 14 | Emphasis on integration into existing health systems and services, particularly for guidelines. More limited reference to the how, including steps such as situational analysis, needs assessment and planning/ budgeting for the necessary infrastructure, resources and capacity development. |
| Monitoring and evaluation | 10 | 13 | A need for M&E frameworks often mentioned but lacking in detail. |
| Wider multisectoral support | 3 | 9 | Minimally mentioned with only one guideline emphasising the need for linkages to other services and systems (social assistance/protection, food, water and sanitation) and two making brief mentions of policies/services outside of the health system. Greater presence in manuals and guidance documents but still low overall and lack detail when included. |
| Organisational capacities | 3 | 6 | Largely missing, except for few examples which included best practice statements for, or needs to align with, emergency preparedness plans or humanitarian response strategies. |

\*MM and HD combined appraisal notes

## DISCUSSION

In this review, we examined global policy content related to the care of vulnerable infants u6m and their mothers. In our discussion, we explore implications of our findings regarding coherence, opportunities, and gaps within and across policies. We consider policy content in relation to policy context, actors, and guidance development processes (94). We identify strategic and practical directions for enhancing policy-directed care guided by national policymaker interests (16).

### Policy content

#### Characteristics

Our review revealed a global policy landscape that is rich and diverse but also highly fragmented.Policies span multiple disciplines and development processes, resulting in variable content both across and within guidance documents over time. We identified a wide range of vulnerability descriptors that, while seemingly distinct, often refer to overlapping populations. This creates artificial distinctions and risks obscuring opportunities for alignment. For example, a “small vulnerable newborn” may be later identified as “malnourished” or “underweight” is often treated as a different case in policy, despite being the same child clinically. Conversely, terminological similarities can obscure important differences. The 2023 WHO malnutrition guideline, for instance, expanded its scope from severely malnourished infants u6m (9) to infants u6m at risk of poor growth and development (35). However, while the population covered has broadened, some 2013 recommendations—such as those on antibiotic use—were not updated. The existing 2013 recommendation on antibiotics still stands, though it is not transferable to the wider group of at-risk infants now included in the guideline. These inconsistencies undermine policy coherence, reduce content predictability, and hinder accessibility. They create a layer of complexity that risks confusion for implementers, and mask both critical policy gaps and inter-policy potential.

#### Vulnerability of infants

Poor infant growth and low anthropometry were underrepresented as markers of vulnerability in the policies reviewed. Growth and anthropometric indicators were more frequently addressed in nutrition-oriented policy guidance. In most postnatal guidance, LBW is treated primarily as a risk factor for an acute event (birth) rather than the starting point for continued growth monitoring and appraisal. This likely reflects the narrow life-course focus of many postnatal policies, which tend to prioritize the high-risk neonatal period.

Clinically oriented guidance sometimes addressed breastfeeding difficulties but less commonly included growth assessment. Underweight is recommended as a risk criterion in only one WHO guideline (35), is limited to infants under two months in IMCI guidance (13, 66), and is absent from WHO clinical policy documents. These omissions are not aligned with current evidence, which shows that underweight infants u6m—particularly those with comorbidities requiring inpatient care—face substantially higher mortality risk (31). This discrepancy may stem from a prevailing perception that anthropometric indicators are specific to nutrition, rather than being recognized as sensitive predictors of mortality. Addressing this gap may require a conceptual shift—one that reframes anthropometric measures as key indicators of vulnerability across health domains, not just within nutrition. This is necessary to ensure management embeds but extends beyond nutrition interventions.

#### Vulnerability of mothers

Maternal health was commonly considered in policy guidance. However, content was inconsistent and typically framed in relation to infant needs, rather than from the mother’s perspective. There was limited coverage of maternal nutritional status, anaemia, and anthropometry. Postnatal guidelines offered greater detail on maternal components, that may reflect how mothers’ wellbeing is inherent in early antenatal care. However we found, like others have, this tapered off thereafter (95). Where maternal care was embedded as a core component of at-risk infant care (26, 35, 62), implementation details remain weighted towards infant needs. The limited coverage of maternal needs beyond the immediate postnatal period may reflect the typical segregation between adult and child health services. These policy gaps may also reflect broader, systemic shortcomings in providing adequate provision for women’s health and nutrition (96, 97). If so, even if policies improve, implementation requires services that may be inconsistent or non-existent.

These limitations have far-reaching implications, not only for infant health, but also for the rights and wellbeing of women themselves. It is well established that a mother’s physical and mental wellbeing influences her ability to nourish and care for her infant, as well as to maintain her own health. However, care for an at-risk infant draws heavily on a mother’s mental and physical resources. She is a woman who has her own vulnerabilities and needs. Safeguarding and strengthening a mother’s nutrition and health status is essential for delivering responsible, equitable, and effective care (98, 99).

#### Continuity of care

Continuity of care was frequently cited as a guiding principle in the policies reviewed, but it was often not well operationalized. Policy content tended to focus on the management of specific conditions—such as feeding difficulties or acute illness—with much lower coverage of the informational and relational components of person-centred care (100). This limitation may partly reflect the scope of policy guidance itself. For example, WHO guidelines provide evidence-based recommendations on selected aspects of care for a particular population or condition, which may not extend to broader, cross-cutting components of continuity of care.

Policy guidance for vulnerable newborns primarily concentrated on the early postnatal period, with a strong focus on breastfeeding and clinical care. However, there was limited guidance on follow-up into later infancy and early childhood. Infants born small-for-gestational age received minimal policy attention. While some nutrition and health policies mention follow-up for babies born too small or too early, overall policy coverage remains limited. Notably, low birth weight and prematurity are not recognized as risk markers within IMCI guidance (13, 66).

We also observed areas of strong synergy across policy documents. In some cases, these links were explicit, for example, WHO tuberculosis guidelines are directly supported by derivative operational guidance. In others, cross-references were embedded within policy texts - for instance, the 2023 updated WHO malnutrition guidelines cross-refer to neonatal care guidance for very young infants. However, some synergies remained implicit. The MAMI Care Pathway Package, for example, aligns closely with the 2023 malnutrition guideline recommendations but is not directly referenced within them.

These findings highlight missed opportunities for inter-policy alignment to support more coherent and continuous care for vulnerable infants and their mothers. Small vulnerable newborns become small vulnerable babies but this is not recognisable in policy content (5, 101). Realizing this potential will require more intentional integration and bridging of related guidance across sectors and life-course stages.

### Policy processes

Our findings illustrate that policy coherence requires policy connectivity. We found, like others have, that there are practical challenges in applying WHO’s living guidelines approach to more complex fields of care (102). For example, updates to even one recommendation requires cascading revisions across multiple UN and non-UN documents. The living guidelines approach provides a means for responsive updates but so far, has only been applied to some policy guidance, such as COVID (103,104), and there is no official guidance on how to implement it (15). We identified immediate needs in this regard. For example, a recent secondary data analysis identified three “readily assessable” indicators (not-breastfed, LBW/premature, underweight) to inform risk-differentiated care in services (31), yet all three had low coverage across WHO and non-UN guidance. There are also outstanding needs, for example, WHO clinical guidelines only reference WHZ <-3 from the 2013 malnutrition guideline that is now outdated (31). Furthermore, no mechanism exists to systematically identify where inconsistencies and gaps may lie in what already exists — recognising and identifying the extent of a problem is essential to figuring a solution.

### Ways forward

Our findings reflect the inherent complexity of real-world policy and implementation contexts and interactions (105). Improving coherence requires approaches that recognise and embrace this rather than attempt to mute it. The challenge lies in how to inter-play policy, evidence, and practice without furthering complications or heightening resource demands. Working with what exists and considering what is needed, we make several suggestions to strengthen policy coherence most immediately and in the future.

#### Toward a living policy system

Our findings suggest the need for a dynamic, living policy system to improve accessibility, identify synergies across disciplines, and support the alignment of policy processes. We envision an interlinked, continuously updated policy curation mechanism, framed around person-centred continuity of care for vulnerable infants u6m and their mothers. This umbrella framework could help situate diverse policy guidance within the broader context of care continuity, both within and across sectors. Active policy surveillance would enable timely updates in response to emerging evidence and facilitate the dissemination of policy addenda or revisions. Integrated feedback mechanisms could help identify and respond to the priorities and needs of national policymakers, ensuring that global guidance remains relevant and valued. Embedding learning processes within this system could also support the implementation of ‘living guidelines’ for complex care scenarios (15, 102) while contributing to improved evaluation of the impact of normative guidance (106).

Two existing WHO initiatives offer promising models for such an approach. First, the WHO Clinical Care in Crises (CCC) Mobile App development for frontline workers (107). It used an international advisory group to identify and address policy inconsistencies to produce algorithms. Second, the COVID-19 Living Guidelines mechanism—a collaboration between WHO, the MAGIC Evidence Ecosystem Foundation, and *The BMJ*—uses a standing expert panel to produce real-time, living network meta-analyses, with regular updates and alerts (104) hosted on the MAGICApp platform (108). Several UN guidelines we reviewed are already hosted on MAGICApp. However, what remains lacking is a comprehensive, curated collection of interconnected policy content, structured under a unified framework and actively managed as a living system.

#### Handling complexity within policy guidance

Our review suggests that methodological rigour alone does not ensure consistency in guideline outputs (7). The significant variation we observed across WHO guideline content likely reflects attempts to address the “whether, when, and how” of delivering complex care within the constraints of condition- or population-specific guidance. Emerging frameworks such as WHO-INTEGRATE (109) offer valuable tools for managing complexity within the GRADE process (7). The variability we observed across guidance also reflects the varied sources and approaches of many stakeholders in developing content. Broader application of quality assurance frameworks such as AGREE and WHO-INTEGRATE enhance the rigor of both UN and non-UN content, support greater policy coherence, and contribute to methodological development and its evaluation (110, 111).

Implementation guidance developed at the global level will inevitably have limitations when it comes to contextualising to diverse and dynamic settings. National stakeholders are best positioned to adapt global guidance tailoring recommendations to their local population needs and system capacities. Incorporating frameworks like WHO-INTEGRATE into national adaptation processes could help support national decision-making by helping to navigate uncertainty and balance trade-offs. WHO regional and country offices could facilitate this, for example, by deploying roving systems specialists to foster cross-country learning and collaboration. Furthermore, WHO could benefit from systematically learning from country-led adaptations. Insights from these experiences could inform the organization’s own evidence-to-policy methodologies and contribute to institutional impact evaluations of normative guidance development (112).

#### Investing in policy cooperation

Ongoing policy processes at the WHO present a timely opportunity to strengthen interdepartmental collaboration, align outputs, and support national policy development. For example, current guidance development work - spanning malnutrition, care for small and sick newborns, and IMCI - offers a concrete opportunity to promote policy coherence and ensure consistent, aligned content. While inter-policy collaboration within WHO often occurs informally, such ad hoc coordination requires considerable time and effort. To enhance efficiency and impact, an institutionalized mechanism is needed, that mandates and resources interdepartmental coordination as a core function. Given increasingly constrained national and global resources, streamlining processes across disciplines and departments offers clear strategic and economic advantages (113, 114). Leveraging existing tools and evidence represent a strong investment case.

Importantly, WHO is not the sole source of public health guidance. Content produced by non-UN actors can also meet rigorous standards and be highly relevant. Our review identified several opportunities for cross-agency policy cooperation. For instance, the MAMI Care Pathway Package applied AGREE criteria and was developed through expert-peer collaboration. It aligns with WHO’s 2023 malnutrition guidelines, is modelled on and extends the IMCI guidance, and incorporates outpatient follow-up for vulnerable newborns. WHO has recognized it as a “concrete illustration” of how to operationalize recommendations for risk-differentiated care (115) and results from an evaluated randomized controlled trial (RCT) in Ethiopia are expected imminently. However, WHO procedures do not currently provide a clear mechanism for incorporating non-UN guidance into its formal processes (7). WHO ‘guidance on guidelines’ do not include appraisal of existing guidance and tend to assume that implementation guidance is derived from WHO recommendations. Our review found that, in some cases, non-UN guidance supporting WHO recommendations preceded the guideline publication. Updating WHO’s procedures to also consider high-quality non-UN guidance could help address urgent policy gaps, ease the burden on WHO’s own guideline development processes, and reduce duplication of effort.

#### Shared problems may spark collective solutions

Multiple descriptors of vulnerability risk creating artificial distinctions between what are often interconnected challenges. Siloed framing contributes to siloed policies that risk siloed services. Inconsistent definitions and fragmented guidelines have hindered action on LBW within the reproductive health field (116); our review suggests similar consequences are likely in the broader maternal-infant health landscape. Process-generated differences may fuel sectoral competition, national confusion and missed opportunities for integrated care. While agreeing common terms across stakeholders is likely unrealistic, securing shared understanding and pursing common goals is. A clearly defined, shared problem could help consolidate efforts, maximise existing tools, and attract attention and resources that are typically insufficient across health and nutrition domains (117).

### Strengths and limitations

This scoping review focused on global policy guidance and did not assess evidence from trials or interventions. We did not evaluate the risk of bias or the quality of policy documents, as these are beyond the remit of scoping reviews. We excluded enabling policy guidance that lacked sufficient depth for our practice-centred focus. Ethical approval was not required, as we used only publicly available data.

We reviewed only English-language publications. Four regional/national documents were included due to their global relevance (39, 62, 67, 69) but as we did not systematically source these, others may have been missed. We used our professional networks to source policies, which may have introduced selection bias. We did not formally appraise policy context, stakeholders and processes. Stakeholder consultation would have strengthened the review but was not feasible; this remains an important area for future research (21). Future reviews should also seek documents in other languages and investigate the extent of translation of global guidance produced in English. Our own experience, assumptions, and relationships shaped this review, potentially introducing bias, but also adding value. We welcome feedback and encourage others to build on this work.

## Conclusions

We found a rich melting pot of valuable policy guidance relevant to the care of vulnerable infants u6m across sectors and specialities. Global policy guidance spans multiple disciplines, requiring improved transparency, coherence, and collaboration to manage its inherent complexity. There is both need and opportunity to defragment existing policies, develop inter-speciality understanding and secure inter-sectoral policy pathways of care. There are inter-policy synergies to leverage and pressing gaps to address on growth, follow-through care for small vulnerable newborns and maternal nutrition and health. The COVID living guideline mechanism exemplifies the value of policy navigation. It is time to match the same ambition and investment to vulnerable infants and their mothers. National leadership to contextualise global policy content will maximise the potential for positive policy impact. These actions are an economic and equitable imperative for securing the best possible care for vulnerable infants u6m and mothers.

## Data Availability

All policy guidance we including in our review is publicly available. We have cited these documents throughout the manuscript. We have collated them and made the available at an online open access platform to improve accessibilty. We have included weblinks to resource repositories where we accessed content. We have included additional methodological details and data in supplementary information. Our excel worksheets are available on request, as indicated in the manuscript.

https://public.tableau.com/app/profile/mami.global.network/viz/MAMIScopingReview2024/Dashboard1.

## Acknowledgements

We thank Amir Samani (ENN) for early help in cataloguing documents and Eilis Brennan (ENN) for further support in collating, cataloguing and classification of prioritised materials. We thank Louise Day (LSHTM), Martha Mwangome (KEMRI Wellcome Trust, Kenya), Phil James (ENN), Anne Walsh (ENN) and Nicky Dent (ENN) for their feedback on emerging findings.

## ADDITIONAL INFORMATION

S.Appendix 1: PRISMA-ScR Completed Checklist

S.Appendix 2 Reflexivity statement

S.Appendix 3: MAMI Care Pathway Framework

S.Appendix 4: Profile of individuals contacted to source policy documents

S.Appendix 5: Search strategy

S.Appendix 6: Data items

S.Appendix 7: Vulnerability factors (n=24) mapped across 34 policy documents

## Notes

### Competing Interest Statement

The authors have declared no competing interest.

### Funding Statement

Three authors (MM, SV, HD) were funded to undertake this review by the Gates Foundation (INV-042778) and the Department of Foreign Affairs and Trade, Ireland (HQPCR/2023, 2024/ENN) through agreements with the Emergency Nutrition Network (ENN). MM undertook further unfunded work as part of her PhD studentship at the London School of Hygiene and Tropical Medicine (LSHTM). Two authors (MK, TS) received no specific funding for this work.

### Summary of Updates

Correction to typos to version 2 manuscript and S.Appendix 1.

