## Supplementary material for "Invisible pursuit: A scoping review of global policy for continuity of care of vulnerable infants under 6 months and their mothers in low- and middle-income countries": S.Appendix 2

**S.Appendix 2 Reflexivity statement**

The authors are nutrition and health experts with experience developing global and national policy guidance (MM, HD, MK, TS) and conducting scoping reviews (MM, HD, SW, MK, TS).

MM and MK co-conceptualised and have overseen development of the MAMI Care Pathway Package applied in this review. MM and MK co-chair the MAMI Global Network that coordinated its development through peer and expert consultation. Both MM and MK are researchers on the MAMI RISE Research Project to test the MAMI Care Pathway approach in a RCT and process evaluation in Ethiopia.

We relied on our collective expertise to consider what was feasible and useful for practice-informed policy interpretation. We have drawn upon our lived experiences in global policy processes and witnessed effects at national level to suggest constructive ways forward.

To help interpretation of our interpretations of policies, we include details of who conducted which components of the review. In the discussion, we include details of our reasonings for our findings and recommendations. We address our subjectivity in the strengths and limitations section.
