## Supplementary material for "Invisible pursuit: A scoping review of global policy for continuity of care of vulnerable infants under 6 months and their mothers in low- and middle-income countries": S.Appendix 3

**S.Appendix 3: MAMI Care Pathway framework**

**S.Figure 3a: MAMI Care Pathway components embedded within the formal health system**

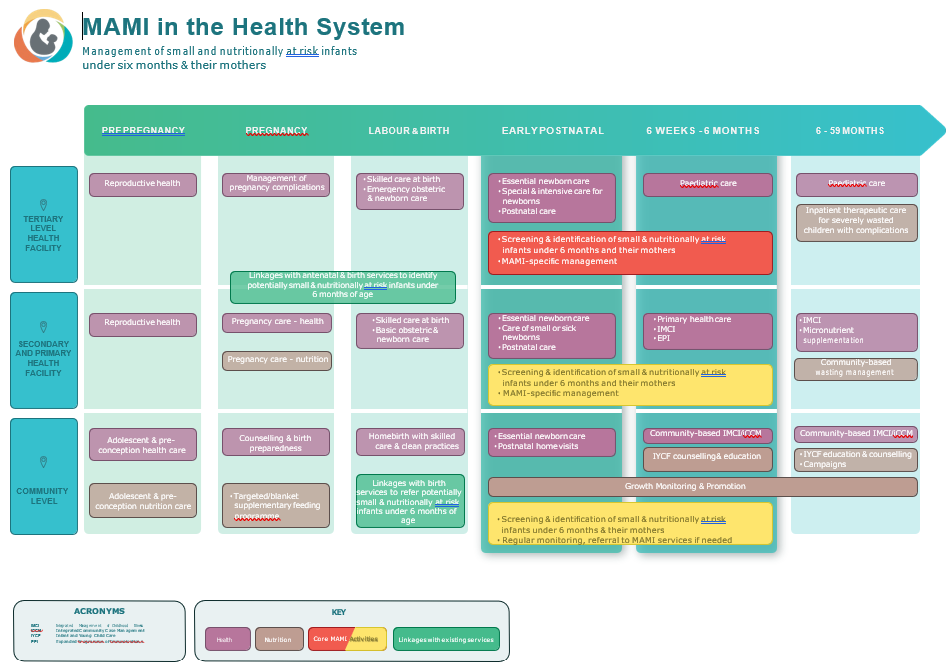

Source: MAMI Global Network, ENN, LSHTM. MAMI Care Pathway Package. v3. 2021. <https://www.ennonline.net/mamicarepathway>

**S. Figure 3b: Mapping ‘who, what, where’ for care provision within existing health services**

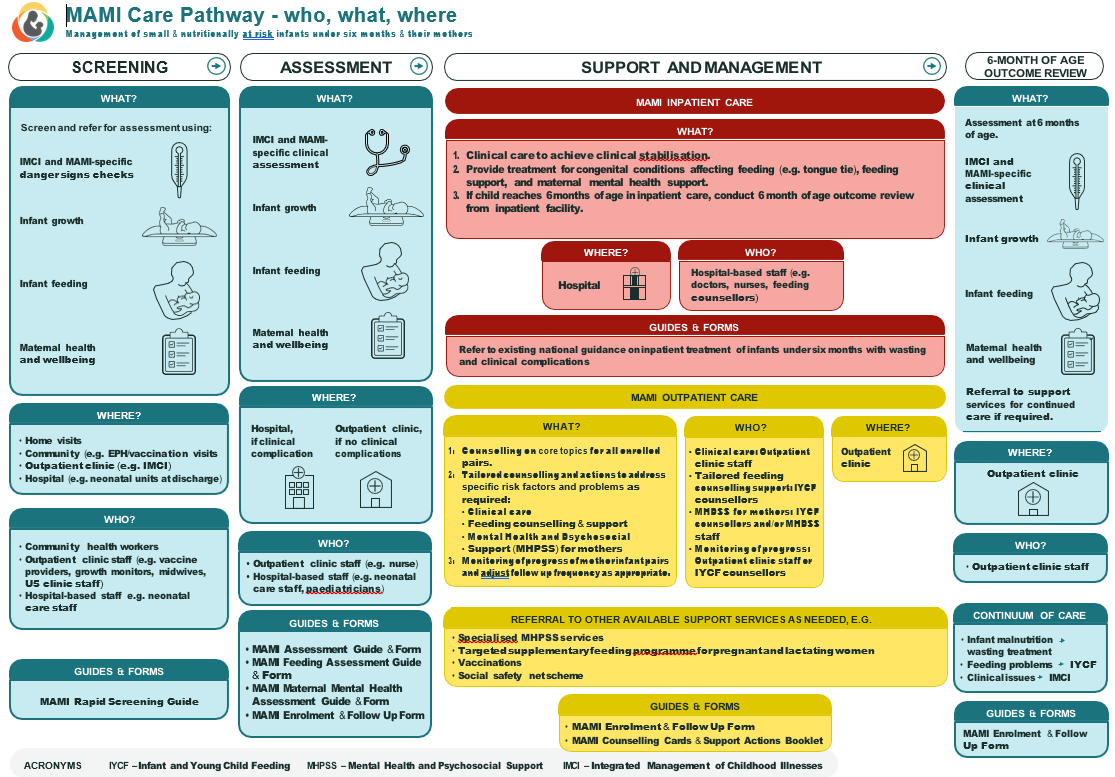

Source: Source: MAMI Global Network, ENN, LSHTM. MAMI Care Pathway Package. v3. 2021. <https://www.ennonline.net/mamicarepathway>
