## Supplementary material for "Invisible pursuit: A scoping review of global policy for continuity of care of vulnerable infants under 6 months and their mothers in low- and middle-income countries": S.Appendix 4

**S.Appendix 4: Profile of individuals contacted to source policy documents**

**Table 4a: Speciality of individuals contacted to source documents**

| **Speciality*** | **n** | **%** |
| --- | --- | --- |
| Nutrition (all) | 18 | 43% |
| - *Nutrition* | *16* | 38% |
| - *Nutrition and health* | *2* | 5% |
| Maternal nutrition | 1 | 2% |
| Child development | 4 | 10% |
| Child/Paediatric health | 13 | 31% |
| Newborn | 10 | 24% |
| Maternal health (all) | 7 | 17% |
| - *Mental health* | 4 | 10% |

*Note: more than one response possible and self-defined categorisation

**Table 4b: Agency affiliation of individuals contacted to source documents**

| **Agency type** | **n** | **%** |
| --- | --- | --- |
| UN | 8 | 19% |
| NGO | 19 | 45% |
| Bilateral donor (one agency) | 3 | 7% |
| Foundation funder (one agency) | 2 | 5% |
| Acaedemic | 8 | 19% |
| Independent | 1 | 2% |
| Technical organisation | 1 | 2% |
| TOTAL | 42 | 100% |
