## Supplementary material for "Invisible pursuit: A scoping review of global policy for continuity of care of vulnerable infants under 6 months and their mothers in low- and middle-income countries": S.Appendix 5

**S.Appendix 5: Search strategy**

**Search terms**

1. newborn* or new-born or neonat* or prematur* or infant* or infancy or baby or babies or p?ediatric*
2. low-birth-weight or LBW or prematur* or small-for-gestational-age or SGA or small-for-age or SFA
3. malnourished or malnutrition or severe malnutrition or severely malnourished or severe acute malnutrition or SAM or moderate* malnutrition or moderately malnourished or moderate acute malnutrition or MAM or acute malnutrition or acutely malnourished or AM or severe wasting or severely wasted or moderate wasting or moderately wasted or wasting or wasted or thin* or stunting or stunted or growth-failure or growth-falter* or poor growth or under-weight or failure-to-thrive or FTT or failure-to-grow or growth delay or delayed growth or nutrition*deficien* or micronutrient* deficien* or nutritionally-at-risk or nutrition disorder* or protein-energy-malnutrition or PEM or development* delay or delayed development or mid-upper-arm-circumference or MUAC or weight-for-length or WFL
4. (1 AND 2) OR (1 AND 3) *[all vulnerable infants]*
5. (mother* or matern*) ADJ2 (health or nutrition or mental-health or reproductive-health or food or social-assistance or social-welfare or nutrition or malnutrition)
6. 4 OR 5 *[all vulnerable infants OR mothers]*
7. policy or policies or guid* or strateg* or manual or framework or plan
8. 6 AND 7 *[all vulnerable infants OR mothers AND guidance]*
7. Screen for identified documents relevant to infants under six months of age at global level
