## Supplementary material for "Invisible pursuit: A scoping review of global policy for continuity of care of vulnerable infants under 6 months and their mothers in low- and middle-income countries": S.Appendix 6

**S.Appendix 6: Data items**

**Phase 1. Appraising key characteristics**

The data set included the following variables:

Publication year;

URL;

Source;

Title;

Type (e.g., action plan, framework, implementation guidance, guideline, manual, report, training guide);

Aim;

Condition (disease, disorder, injury, disability, or profile of risk factors);

Target audience;

Target population (e.g., infant, mother (principal caregiver), both);

Infant’s description (e.g., healthy newborn, healthy infant, vulnerable newborn, preterm infant (PT), low birth weight infant (LBW), small, sick, (non)breastfed, wasted/acutely malnourished, underweight, stunted, growth faltering, at risk of poor growth and development, feeding problem, excessive crying, disability, acute or chronic illness);

Mother’s description (e.g., healthy, sick, mental health issue, malnourished, adolescent, prenatal, perinatal, postnatal issues, (not)lactating, absent, multipara, primipara, absent, dead);

Sector (e.g., child health, child nutrition, child development, maternal physical health, maternal mental health, maternal nutrition, reproductive health, newborn/neonatal health);

Level of care (e.g., tertiary, secondary, primary or community care)

Care service (e.g., maternity, child health, child nutrition units); level of care (e.g., inpatient, outpatient, community care);

- Age timeline covered;

Infant risks covered (e.g., LWB, PT, SGA, growth faltering (recent weight loss, poor growth, low WAZ, low MUAC), feeding/metabolic problem, excessive crying, disability, acute or chronic illness);

Maternal risks covered (e.g., nutritional risk, impaired BF, physical health, MMH, multipara, primipara, <18 year, absent, dead);

Interventions (e.g., active case finding or screening, health assessment or IMNCI, breastfeeding (BF) assessment, non-BF feeding assessment, BF support, non-BF feeding support, clinical care, nurturing care, early childhood development (ECD), crying and sleep counselling, mental health counselling, social support, follow-up visits, home visits, family involvement, continuity of care);

**Phase 2. Describing vulnerability of infants u6m and their mothers**

The data set included the following variables of vulnerability:

Poor birth outcome

Small newborn:

Low birth weight (LBW) <2.5 kg; LBW <2.0 kg; very LBW <1.5 kg; extremely LBW <1.0 kg

Preterm (PT) <37 weeks gestation; very PT <33 weeks; extremely PT <28 weeks

Small for gestational age (SGA) (birth weight <10th percentile for gestational age)

Sick newborn:

Birth trauma or birth complications

Congenital illness (e.g., congenital heart disease, HIV, TB)

Disability and/or congenital abnormality (e.g., tongue-tie, cleft palate)

Macrocephaly head circumference (HC) (HC-for-age z-score >+3 or rapid crossing z lines (+1 or more z-score change in 2 months))

Microcephaly (HC-for-age z-score ≤-3)

Morbidity related to prematurity

Low infant anthropometry (including nutritional oedema)

Nutritional oedema

WAZ <-2; WAZ <-3

WLZ <-2; WLZ <-3

MUAC <115 mm for infant 6 weeks-<6months

Poor growth based on sequential measures of ponderal growth

Recent weight loss (decreasing weight; downward crossing growth lines)

No weight gain on two consecutive measurements (stationary weight)

Insufficient weight gain (flat WAZ or WLZ growth; less than 500g/kg/month)

Insufficient weight gain for preterm (less than 18g/kg/day and 0.9 cm/week in head circumference)

Risk factors for poor growth and development

Infant's health and feeding risk factors:

Infant breastfeeding (BF) difficulties (e.g., attachment, suckling reflex, refusal, intolerance)

Infant history of hospitalisation

Infant IMCI danger sign or sign of acute medical problem

Infant medical problem needing mid/long term care

Infant mental health (e.g., excessive crying)

Infant neurodevelopment concerns

Non-BF infants (e.g., unsafe preparation and use of BMS, access to BMS)

Severe ill infant and no referral possible

Mother's health, feeding, nutrition, and social risk factors:

Lack of birth spacing

Mother adolescent

Mother dead or absent

Mother's BF concerns (e.g., attachment, positioning, perceived breastmilk insufficiency mixed feeding, other ineffective feeding/time, frequency)

Mother's birth complication

Mother's MUAC

Mother's physical health (e.g., TB, HIV, disability)

Mother's social or contextual factors affecting with care and feeding (other)

Mother’s anaemia

Mother’s mental health

Multipara

Primipara

**Phase 3. Appraising guidance on continuity of care**

The data set included the following variables:

Condition: How is the condition or risk profile described (e.g., health problem, disease, disorder, injury, disability)?

Care across time, services and levels of care: Is care being provided across services, levels of care and time (e.g., assisting referral, providing follow-up post exit/discharge, connecting to follow-on services across health, nutrition, and social services)?

Integrated care pathway: Does care cover assessing, classifying and/or acting on the condition, including monitoring individual progress and outcomes?

Comprehensive person-centred care: Is care organised around the health needs and expectations of the person rather than on disease (including a comprehensive assessment of a person’s needs and building individual resilience; involving a multidisciplinary team)?

Early childhood development (ECD): Is ECD incorporated into care?

Mother (principal caregiver), father and family support: Is the mother, father, and family engaged in care?

Community participation: Is the community sensitised (aware) and involved in the provision and organisation of care?

Embeddedness (mainstreaming in routine care): Are practices incorporated into everyday work building upon existing services (including practical re-organisation of care and staff with new roles and responsibilities; avoiding duplicative actions or vertical, disconnected service delivery)?

Local health system support: Does the policy guidance include strengthening capacities of the local health system (e.g., on governance, finances, information system, health workforce, supplies and technology, and service delivery, community participation)?

Monitoring and Evaluation (M&E): Is M&E for quality improvement comprehensively covered?

Wider multisectoral support: Are there considerations for socio-economic support and assistance (e.g., cash, food, income generation, maternity leave, childcare for working mothers)?

Organisational capacities, including resilience: Are there considerations given for developing or strengthening organisational capacities so they can better respond to (un)expected changes and absorb shocks?
