## Supplementary material for "Invisible pursuit: A scoping review of global policy for continuity of care of vulnerable infants under 6 months and their mothers in low- and middle-income countries": S.Appendix 7

**S.Appendix 7: Vulnerability factors (n=28) mapped across 34 policy documents**

| **Vulnerability**  **Factors:** | | **Poor birth outcomes** | | | | | | | **Low anthropometry or poor growth** | | | **Risk factors related to the infant** | | | | | | **Risk factors related to the mother** | | | | | | | | | | | |
| --- | --- | --- | --- | --- | --- | --- | --- | --- | --- | --- | --- | --- | --- | --- | --- | --- | --- | --- | --- | --- | --- | --- | --- | --- | --- | --- | --- | --- | --- |
| **Title of document** | **Condition** | Congenital illness (incl. HIV, TB) | Low birth weight | Preterm | Disability or congenital abnormality | Preterm morbidity | Birth trauma or complications | Small for gestational age | Poor growth | Low anthropometry | Nutritional oedema | Breastfeeding difficulties | Illness | Not breastfed | Neurodevelopment concerns | Hospitalisation history | Mental health | Physical health | Breastfeeding conditions | Mental health | Social or contextual factors | Birth complication | Adolescent | Multipara | Absent or died | Anaemia | Primipara | Birth spacing | Low MUAC |
| **Guideline documents (blue)** |  |  |  |  |  |  |  |  |  |  |  |  |  |  |  |  |  |  |  |  |  |  |  |  |  |  |  |  |  |
| *2023 WHO Guideline on the prevention and management of wasting and nutritional oedema in infants and children under 5 years* | Poor growth and development | ✔️ | ✔️ | ✔️ | ✔️ | - | - | ✔️ | ✔️ | ✔️ | ✔️ | ✔️ | ✔️ | ✔️ | ✔️ | ✔️ | - | ✔️ | - | ✔️ | ✔️ | ✔️ | ✔️ | - | ✔️ | - | - | - | - |
| *2022 WHO Consolidated guidelines on tuberculosis in children and adolescents* | Tuberculosis | ✔️ | - | - | - | - | - | - | ✔️ | ✔️ | ✔️ | - | ✔️ | - | - | - | - | ✔️ | - | - | - | - | - | - | - | - | - | - | - |
| *2022 WHO Recommendations for care of the preterm or low-birth-weight infant* | Preterm, LBW |  | ✔️ | ✔️ | - | - | ✔️ |  | - | - | - | ✔️ | ✔️ | ✔️ | - | - | - | - | ✔️ | - | - | - | - | - | - | - | - | - | - |
| *2021 WHO-PAHO Evidence-based clinical practice guidelines for the follow-up of at-risk neonates* | Newborn care | ✔️ | ✔️ | ✔️ | ✔️ | ✔️ | - | - | ✔️ |  |  | ✔️ |  |  | ✔️ |  |  |  | ✔️ | ✔️ | ✔️ | - | - | - | - | - | - | - | - |
| *2021 WHO Guideline: Infant feeding in areas of Zika virus transmission* | Zika virus | ✔️ | - | - | - | - | - | - | - | - | - | ✔️ | - | - | - | - | - | ✔️ | ✔️ | - | - | - | - | - | - | - | - | - | - |
| *2021 WHO Consolidated guidelines on HIV prevention, testing, treatment, service delivery and monitoring* | HIV | ✔️ | - | - | - | - | - | - | - | - | - | - | - | - | - | - | - | ✔️ | - | - | - | - | - | - | - | - | - | - | - |
| *2020 WHO Improving early childhood development* | ECD | - | - | - | - | - | - | - | - | - | - | - | - | - | - | - | - | - | - | ✔️ | - | - | - | - | - | - | - | - | - |
| *2018 WHO Guideline: Counselling of women to improve breastfeeding practices* | Breastfeeding | - | - | - | - | - | - | - | - | - | - | ✔️ | - | - | - | - | - | - | ✔️ | ✔️ | ✔️ | ✔️ | ✔️ | ✔️ | - | - | ✔️ | - | - |
| *2016 WHO Paediatric emergency triage, assessment and treatment (ETAT)* | Critically ill child | ✔️ |  | ✔️ | - | - | - | - | - | - | - | - | ✔️ | - | - | - | - | - | - | - | - | - | - | - | - | - | - | - | - |
| *2015 WHO Recommendations on interventions to improve preterm birth outcomes* | Preterm | ✔️ | ✔️ | ✔️ | ✔️ | ✔️ | ✔️ | ✔️ | - | - | - | - | ✔️ | - | - | - | - | ✔️ | - | - | - | ✔️ | - | - | ✔️ | - | - | - | - |
| *2015 WHO Guideline: Managing possible serious bacterial infection in young infants when referral is not feasible* | Ill child | - | - | - | - | - | ✔️ | - | - | - | - | ✔️ | ✔️ | - | - | - | - | - | - | - | - | - | - | - | - | - | - | - | - |
| *2012 WHO Recommendations for management of common childhood conditions* | Ill child | - | - | ✔️ | ✔️ | ✔️ | ✔️ | ✔️ |  | ✔️ | ✔️ | ✔️ | ✔️ | ✔️ | ✔️ | - | - | ✔️ | ✔️ | - | - | - | - | - | - | - | - | - | - |
| *2012 WHO Guidelines on basic newborn resuscitation* | Newborn resuscitation | - | - | ✔️ | - | - | ✔️ | - | - | - | - | - | - | - | - | - | - | - | - | - | - | - | - | - | - | - | - | - | - |
| **Guidance documents: guides (green) and manuals (yellow)** |  |  |  |  |  |  |  |  |  |  |  |  |  |  |  |  |  |  |  |  |  |  |  |  |  |  |  |  |  |
| *2022 WHO Guide for integration of perinatal mental health in maternal and child health services* | Perinatal mental health | - | - | - | - | - | - | - | - | - | - | - | - | - | - | - | - | ✔️ | ✔️ | ✔️ | ✔️ | ✔️ | ✔️ | - | - | - | - | - | - |
| *2022 UNICEF, University of Pretoria Feeding preterm and low-birthweight newborns* | Preterm, LBW | ✔️ | ✔️ | ✔️ | ✔️ | ✔️ | - | - | - | - | - | ✔️ | - | - | - | - | - | ✔️ | ✔️ | ✔️ | ✔️ | - | - | ✔️ | - | - | - | - | - |
| *2022 WHO-Europe Pocketbook of primary health care for children and adolescents* | Ill child | ✔️ | ✔️ | ✔️ | ✔️ | ✔️ | ✔️ | - | - | ✔️ | ✔️ | ✔️ | ✔️ | ✔️ | ✔️ | ✔️ | ✔️ | ✔️ | ✔️ | ✔️ | ✔️ | ✔️ | - | - | ✔️ | ✔️ | - | - | - |
| *2022 SPOON Identifying feeding difficulties in infants – guidelines for healthcare professionals* | Feeding difficulties | ✔️ | - | - | ✔️ | - | - | - | ✔️ | - | - | ✔️ | - | - | - | - | - | - | - | - | - | - | - | - | - | - | - | - | - |
| *2022 SPOON Screening children for feeding difficulties* | Feeding difficulties | ✔️ | - | - | ✔️ | - | - | - | ✔️ | - | - | ✔️ |  | ✔️ | ✔️ | - | - | - | - | - | - | - | - | - | - | - | - | - | - |
| *2022 UNICEF Integrating early detection and treatment of child wasting into routine primary health care services* | Wasting | - | - | - | - | - | - | - | - | - | - | - | - | - | - | - | - | - | - | - | - | - | - | - | - | - | - | - | - |
| *2022 WHO Operational handbook on tuberculosis. Module 5: Management of tuberculosis in children and adolescents* | Tuberculosis | ✔️ | - | - | - | - | - | - | ✔️ | ✔️ | ✔️ | - | - | - | - | - | - | ✔️ | - | - | - | - | - | - | - | - | - | - | - |
| *2021 ENN, LSHTM MAMI Care Pathway Package, Version 3 (guiding framework)* | Small and nutritionally at-risk infants and their mother | ✔️ | ✔️ | ✔️ | ✔️ | - | - | ✔️ | ✔️ | ✔️ | ✔️ | ✔️ | ✔️ | ✔️ | ✔️ | - | ✔️ | ✔️ | ✔️ | ✔️ | ✔️ | - | ✔️ | ✔️ | ✔️ | - | ✔️ | - | ✔️ |
| *2020 WHO, UNICEF The Baby Friendly Hospital Initiative for small, sick and preterm newborns* | Breastfeeding | - | ✔️ | ✔️ | ✔️ | ✔️ | - | ✔️ | ✔️ | - | - | ✔️ | ✔️ | ✔️ | ✔️ | - | - | - | ✔️ | - | - | - | - | - | - | - | - | - | - |
| *2020 Partners in Health, UNICEF Early Childhood Development Support for High-Risk Infants* | Preterm, LBW | ✔️ | ✔️ | ✔️ | ✔️ | ✔️ | ✔️ | - | ✔️ | ✔️ | - | ✔️ | ✔️ | ✔️ | ✔️ | ✔️ | ✔️ | ✔️ | - | ✔️ | ✔️ | - | - | ✔️ | ✔️ | - | - | - | ✔️ |
| *2019 WHO Integrated Management of Childhood Illness of the sick young infant age up to 2 months. Chart booklet* | Ill child | ✔️ | ✔️ | - | - | - | - | - | - | ✔️ | ✔️ | ✔️ | ✔️ | ✔️ | - | - | - | - | ✔️ | - | - | - | - | - | - | - | - | - | - |
| *2018 IASC Field Manual on Reproductive Health in Humanitarian Settings* | Reproductive health | ✔️ | ✔️ | ✔️ | - | ✔️ | ✔️ | - | - | - | - | ✔️ | - | ✔️ | ✔️ | - | - | ✔️ | ✔️ | ✔️ | ✔️ | ✔️ | ✔️ | - | ✔️ | - | - | - | - |
| *2016 WHO Oxygen therapy for children* | Ill child | ✔️ | ✔️ | ✔️ | - | - | - | - | - | - | - | - | ✔️ | - | - | - | - | - | - | - | - | - | - | - | - | - | - | - | - |
| *2015 WHO, UNICEF, USAID Caring for newborns and children in the community: planning handbook for programme managers and planners* | Newborn care | ✔️ | ✔️ | - | - | - | - | - | - | - | - | - | - | - | - | - | - | ✔️ | - | - | - | - | - | - | - | - | - | - | - |
| *2014 WHO-Europe Hospital care for mothers and newborn babies: quality assessment and improvement tool* | Newborn care | ✔️ | ✔️ | ✔️ | - | ✔️ | ✔️ | ✔️ | ✔️ | - | - | ✔️ | ✔️ | ✔️ | - | ✔️ | - | ✔️ | ✔️ | ✔️ | ✔️ | ✔️ | ✔️ | - | - | - | - | - | - |
| *2014 WHO Integrated management of childhood illness – Chart booklet* | Ill child | ✔️ | ✔️ | ✔️ | ✔️ | ✔️ | ✔️ | - | ✔️ | - | ✔️ | ✔️ | ✔️ | ✔️ | ✔️ | - | - | ✔️ | ✔️ | ✔️ | ✔️ | - | - | - | - | ✔️ | - | ✔️ | - |
| *2014 Christian Blind Mission (CBM) Recognising impairments at birth* | Newborn care | - | - | - | ✔️ | - | - | - | - | - | - | - | - | - | ✔️ | - | - | ✔️ | - | - | - | - | - | - | - | ✔️ | - | - | - |
| *2013 WHO Pocketbook of hospital care for children* | Ill child | ✔️ | ✔️ | ✔️ | ✔️ | ✔️ | - | - | ✔️ | ✔️ | ✔️ | ✔️ | ✔️ | ✔️ | ✔️ | ✔️ |  | ✔️ | ✔️ | ✔️ | ✔️ | - | - | ✔️ | - | - | - | - | - |
| *2012 CBM Cerebral Palsy* | Cerebral palsy | - | - | - | ✔️ | ✔️ | ✔️ | - | - | - | - | - | - | - | ✔️ | - | - | ✔️ | - | ✔️ | - | - | - | - | - | - | - | ✔️ | - |
| *2003 WHO Kangaroo mother care: a practical guide* | Newborn care | - | ✔️ | ✔️ | - | ✔️ | - | ✔️ | ✔️ | - | - | ✔️ | ✔️ | ✔️ | - | - | - | ✔️ | ✔️ |  | ✔️ | ✔️ | ✔️ | ✔️ | ✔️ | - | - | - | - |
| *2003 WHO Managing newborn problems* | Newborn care | ✔️ | ✔️ | ✔️ | ✔️ | ✔️ | ✔️ | - | ✔️ | - | - | ✔️ | ✔️ | ✔️ | ✔️ | ✔️ | - | ✔️ | ✔️ | ✔️ | ✔️ | ✔️ | ✔️ | - | - | - | ✔️ | - | - |

✔️= vulnerability factor covered; - = vulnerability factor not covered; ECD= early childhood development; LBW= low birth weight; n/a= not applicable; PT=preterm.
